# Maternal and perinatal outcomes of early- versus late-onset Preeclampsia at a tertiary hospital in Bangladesh: A retrospective cohort study

**DOI:** 10.1101/2025.10.15.25337483

**Authors:** Taukir Tanjim, Saleh Haider, Hafsa Hossain, Rajib Biswas, Dilruba Zeba

## Abstract

**Background:** Pre-eclampsia (PE) is a significant cause of maternal and perinatal morbidity and mortality worldwide. The clinical course and severity of PE can vary depending on the gestational age at onset. However, limited data from low- and middle-income countries (LMICs) including Bangladesh stratify outcomes by onset timing, hindering context-specific management. This study aimed to differentiate maternal and perinatal outcomes associated with early-onset PE (EO-PE, <34 weeks) versus late-onset PE (LO-PE, ≥34 weeks).

**Methods:** A retrospective cohort study was conducted at a tertiary-level hospital in Bangladesh from March to August 2023. Data were extracted from hospital records and supplemented through structured telephone interviews for women diagnosed with PE. Associations between PE onset timing and adverse outcomes were analysed using simple and multivariable logistic regression.

**Results:** Out of 3785 admitted patients, 11.28% (n=427) was diagnosed with PE. Of them, 255 PE patients were included in the study (EO-PE: n=121; LO-PE: n=134). EO-PE was associated with more adverse perinatal outcomes compared to LO-PE, including higher rates of prematurity, low birth weight, and increased need for neonatal hospital admission. Maternal complications such as eclampsia (31% vs 19%) and placental abruption (20% vs 10%) were more frequent in the EO-PE group. In multivariable regression, EO-PE was not independently associated with maternal adverse outcomes but showed significantly higher odds of stillbirth (aOR: 17.7, 95% CI: 6.15-66.7, p<0.001) and neonatal adverse outcomes (aOR: 34.6, 95% CI: 13.7-106, p<0.001).

**Conclusions:** Early-onset PE is associated with substantially worse adverse perinatal outcomes compared to late-onset PE. Onset-based classification, early screening, and targeted management strategies are recommended to reduce the burden of pre-eclampsia in Bangladesh and similar LMIC settings.

## 1. Introduction

Pregnancy, while a transformative phase in a woman’s life, can be complicated by conditions that pose significant risks to both mother and child (1). Hypertensive disorders of pregnancy, particularly pre-eclampsia (PE), are major contributors to maternal and perinatal morbidity and mortality globally (2). PE is typically characterized by new-onset hypertension after 20 weeks of gestation accompanied by proteinuria or other evidence of maternal end-organ or uteroplacental dysfunction (3). It affects an estimated 2% to 10% of pregnancies worldwide, with 20% of the total global PE patients belong to the underdeveloped nations (2, 4). The burden of PE is disproportionately high in developing countries, which may range from 2% to 17% due to challenges in healthcare infrastructure and management of complications (5, 6).

Bangladesh has made significant progress in reducing overall maternal mortality in recent decades. However, the prevalence of PE is 14.4% in Bangladesh which is much higher compared to the global percentage (7). According to Bangladesh Maternal Mortality and Health Care Surveys (BMMSs) 2016, the preeclampsia/eclampsia-specific mortality ratio was 46 per 100,000 live births, (8). Moreover, 9% of stillbirths and 7% of early neonatal deaths in Bangladesh was attributed to eclampsia, the severe progression of PE (9). PE remains a formidable challenge, accounting for nearly one-fourth of all maternal deaths and ranking as the second most common direct cause of maternal mortality in Bangladesh (8).

The classification of PE into early-onset (EO-PE, <34 weeks) and late-onset (LO-PE, ≥34 weeks) has important prognostic implications (10, 11). EO-PE is typically driven by defective deep placentation and inadequate spiral artery remodeling, resulting in more severe uteroplacental insufficiency and poorer outcomes (12, 13). Understanding these distinctions is critical for risk stratification and targeted interventions, and optimizing resource allocation – particularly in LMIC settings where health infrastructure is constrained.

Despite the recognized importance of this classification, there is a relative scarcity of studies directly comparing maternal and fetal outcomes between EO-PE and LO-PE groups within the context of Bangladesh and other developing countries. Existing local data do not stratify by onset timing, limiting the ability to tailor management strategies effectively. A better understanding of PE, including its varying clinical presentations and outcomes based on the timing of onset, is essential for improving maternal and perinatal health. Therefore, this retrospective cohort study aimed to compare the maternal, fetal, and neonatal outcomes of early versus late-onset PE among women managed at a tertiary referral hospital in Faridpur, Bangladesh.

## 2. Materials and methods

### 2.1 Study design and setting

This retrospective cohort study was conducted at the Department of Obstetrics and Gynecology, Faridpur Medical College Hospital (FMCH) in Faridpur, Bangladesh. FMCH is a tertiary level hospital serving as a major referral center for public and private hospitals in Faridpur and adjacent districts including Rajbari, Gopalganj, Magura, and Madaripur.

### 2.2 Study population and sampling

We reviewed hospital admission records of all pregnant women admitted and delivered at FMCH between March 2023 and August 2023. Women diagnosed with PE were considered eligible.

Pre-eclampsia was diagnosed by the treating obstetricians in accordance with the institutional clinical practice based on the American College of Obstetricians and Gynecologists (ACOG) recommendations. The diagnostic criteria included new-onset hypertension after 20 weeks of gestation (systolic blood pressure ≥140 mmHg and/or diastolic blood pressure ≥90 mmHg on two occasions at least 4 hours apart in a previously normotensive woman) with either proteinuria (≥300 mg in a 24-hour urine collection, a protein-to-creatinine ratio ≥0.3, or a urine dipstick reading of ≥2+) or evidence of maternal end-organ dysfunction in the absence of proteinuria (3).

Patients whose pregnancy with PE was terminated before the threshold of local fetal viability (defined as <28 weeks of gestation – consistent with institutional practice in Bangladeshi facilities) and those without significant hospital record data or unavailable contact information were excluded. All eligible patients presenting during the six-month study period were enrolled, consistent with a census sampling approach in which the entire accessible population constitutes the sample rather than a probabilistic subset. This eliminated the need for a pre-specified power calculation, as the sample size was determined by the totality of eligible cases at FMCH between March and August 2023 rather than by a power target.

### 2.3 Data collection

Socio-demographic information and other clinical history such as family history and ANC utilization were collected through telephone interviews using a structured questionnaire. Obstetric, delivery, and neonatal data were extracted from hospital medical records. The study abstracted the documented diagnosis of preeclampsia from hospital medical records; superimposed preeclampsia and severe features were not classified separately. No independent verification of diagnoses was performed, and all diagnoses were accepted as recorded.

### 2.4 Variables

The primary exposure variable was the type of preeclampsia, categorized as early-onset preeclampsia (EO-PE) and late-onset preeclampsia (LO-PE). Onset of preeclampsia was defined as the time of first clinical diagnosis documented in the medical records. Early-onset preeclampsia was defined as diagnosis before 34 weeks of gestation, and late-onset preeclampsia as diagnosis at or beyond 34 weeks (10). Gestational age was determined using the best available obstetric estimate, based on a hierarchical approach prioritizing first-trimester ultrasound where available, followed by last menstrual period and then second- and third-trimester ultrasound assessments.

We defined three principal outcome variables for this study: maternal adverse outcome, fetal adverse outcome and neonatal adverse outcome. The maternal adverse outcome composite included the occurrence of any of the following: eclampsia, abruptio placentae, HELLP syndrome, postpartum hemorrhage, premature rupture of membrane, acute kidney injury, pulmonary edema, cerebrovascular accidents or maternal mortality. Fetal adverse outcome was defined as stillbirth. The neonatal adverse outcome composite included one or more of the following: prematurity, low birth weight, intrauterine growth retardation (IUGR), congenital anomaly, an APGAR score of less than 7 at 5-minute, hospital admission or neonatal mortality. Maternal and fetal adverse outcomes were assessed among all pregnancies. Neonatal adverse outcome was assessed only among livebirth cases.

### 2.5 Data management and statistical analysis

Missing data were not a concern in this dataset. Cases with incomplete hospital records were excluded prior to enrolment, ensuring that all included participants had complete clinical documentation. For outcome variables, the absence of a documented complication in the structured hospital record was coded as the condition being absent (binary present/absent ascertainment). Sociodemographic variables collected via telephone interview were fully obtained from all participants who agreed to participate in the study, with no further missing values recorded.

Statistical analysis was performed using STATA version 14.2 (StataCorp, College Station, Texas, USA). Categorical data were presented as frequencies and percentages, and comparisons between EO-PE and LO-PE groups were made using the Chi-square test or Fisher’s exact test where appropriate. Continuous data were summarized as mean ± standard deviation and compared using independent samples t-test. Logistic regression was used to assess the association between PE onset and adverse outcomes. Simple logistic regression was performed first for each outcome to estimate unadjusted odds ratios (OR) with 95% confidence intervals (CI). Multivariable logistic regression was then performed to estimate adjusted odds ratios (aOR), with covariates selected a priori based on clinical relevance and evidence from prior literature. Multicollinearity was assessed across all three multivariable models for maternal, fetal and neonatal outcomes using the generalised variance inflation factor (GVIF). All values were below 2.0 across the maternal (range: 1.04–1.43), fetal (range: 1.00–1.48), and neonatal (range: 1.13–1.57) models, indicating no evidence of multicollinearity among the covariates included in the models. A p-value <0.05 was considered statistically significant.

### 2.6 Ethical considerations

Ethical approval for this study was obtained from the Institutional Review Board (IRB) of Faridpur Medical College Hospital, which does not issue approval numbers for non-clinical trials. Informed verbal consent was obtained from participants prior to telephone interviews. All data collected were anonymized prior to analysis.

## 3. Results

### 3.1 Participant recruitment and baseline characteristics

During the six-month study period, a total of 3785 pregnant women were admitted at the Department of Gynaecology and Obstetrics, FMCH, Faridpur. Among them, 427 (11.28%) mothers were diagnosed with PE. 89 cases were excluded due to the unavailability of significant clinical records, and an additional 41 cases were excluded as our data collectors were unable to contact them. From the eligible cases, another 42 women declined consent. Finally, 255 women with PE agreed to participate, and were included in the study. This final sample comprised 121 women in the EO-PE group and 134 women in the LO-PE group. (Figure-1)

**Figure-1:**
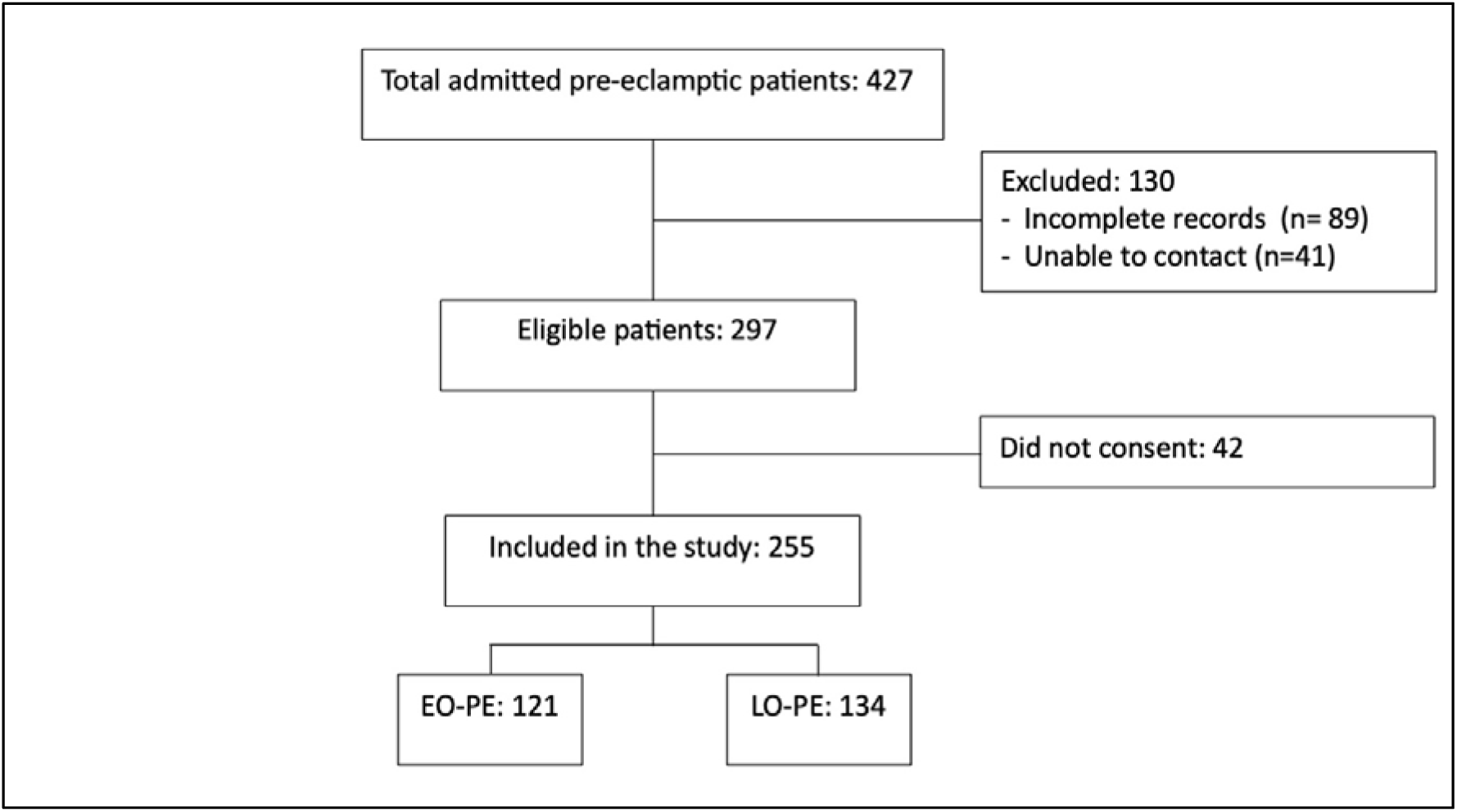
Flowchart of study participant selection

To evaluate potential selection bias, available hospital record data were compared between included (n=255) and excluded (n=172) patients for the variables retrievable from admission registers. No statistically significant differences were observed between the two groups in age (p=0.421), gravidity (p=0.703), or PE onset type (p=0.491).

The mean age of the patients in the final sample was 26.4 ± 6 years, with women in the EO-PE group being older in age compared to the LO-PE group (27.8 ± 5.6 years vs 25.1 ± 5.9 years, p<0.001). While the majority of participants had attained only up to primary level education, the educational attainment was higher in the LO-PE group (p=0.016). Over 80% of participants were homemakers. However, being a student was significantly more common in the LO-PE group compared to the EO-PE group (15.67% vs. 8.26%, p=0.03). About 95% of participants were Muslim, and about two-thirds resided in rural areas, with no difference between groups. (Table-1)

**Table-1:** Comparison of socio-demographic factors between EO-PE & LO-PE groups.

| Characteristic | Early Onset PE<br>N = 121<sup>a</sup> | Late Onset PE<br>N = 134<sup>a</sup> | Overall<br>N = 255<sup>a</sup> | p-value<sup>b</sup> |
| --- | --- | --- | --- | --- |
| **Age** | 27.86 ± 5.64 | 25.11 ± 5.99 | 26.42 ± 5.98 | <0.001 |
| **Living Area** |  |  |  | 0.840 |
| Rural | 88 (72.73%) | 100 (74.63%) | 188 (73.73%) |  |
| Urban | 33 (27.27%) | 34 (25.37%) | 67 (26.27%) |  |
| **Education** |  |  |  | 0.016 |
| Illiterate | 11 (9.09%) | 9 (6.72%) | 20 (7.84%) |  |
| Able to read/write | 18 (14.88%) | 6 (4.48%) | 24 (9.41%) |  |
| Primary | 45 (37.19%) | 44 (32.84%) | 89 (34.90%) |  |
| Secondary | 16 (13.22%) | 30 (22.39%) | 46 (18.04%) |  |
| Undergraduate | 23 (19.01%) | 27 (20.15%) | 50 (19.61%) |  |
| Graduate & above | 8 (6.61%) | 18 (13.43%) | 26 (10.20%) |  |
| **Occupation** |  |  |  | 0.030 |
| Homemaker | 101 (83.47%) | 105 (78.36%) | 206 (80.78%) |  |
| Business | 8 (6.61%) | 2 (1.49%) | 10 (3.92%) |  |
| Student | 10 (8.26%) | 21 (15.67%) | 31 (12.16%) |  |
| Service holder | 2 (1.65%) | 6 (4.48%) | 8 (3.14%) |  |
| **Religion** |  |  |  | 0.637 |
| Muslim | 113 (93.39%) | 128 (95.52%) | 241 (94.51%) |  |
| Hindu | 8 (6.61%) | 6 (4.48%) | 14 (5.49%) |  |

<sup>a</sup> Mean ± SD; n (%)<sup>b</sup> Welch Two Sample t-test; Pearson's Chi-squared test; Fisher's exact test

### 3.2 Maternal clinical history and ANC

A significantly higher proportion of the EO-PE women were multigravid compared to the LO-PE group (76.03% vs. 47.76%, p<0.001). Pre-existing chronic hypertension (14.88% vs 5.97%, p=0.032) and hypothyroidism (14.05% vs. 1.49%, p<0.001) had significantly higher prevalence in the EO-PE group as well. There were no significant differences between groups in the prevalence of gestational diabetes mellitus or a family history of hypertension. Similarly, no significant difference was observed in the history of PE in previous pregnancies among multigravida women (17.39% vs. 12.50%, p=0.544). Regarding ANC, more than half of all participants did not complete the minimum recommended four ANC visits. There was no significant difference in achieving four or more ANC visits between the LO-PE group and the EO-PE group (44.78% vs. 35.54%, p=0.170). (Table-2)

**Table-2:** Comparison of clinical characteristics between the EO-PE and LO-PE groups.

| Characteristic | Early Onset<br>PE<br>N = 121<sup>a</sup> | Late Onset<br>PE<br>N = 134<sup>a</sup> | Overall<br>N = 255<sup>a</sup> | p-value<sup>b</sup> |
| --- | --- | --- | --- | --- |
| Primigravida | 29 (23.97%) | 70 (52.24%) | 99<br>(38.82%) | <0.001 |
| Gestational Diabetes Mellitus | 8 (6.61%) | 8 (5.97%) | 16 (6.27%) | >0.999 |
| Hypothyroidism | 17 (14.05%) | 2 (1.49%) | 19 (7.45%) | <0.001 |
| Chronic Hypertension | 18 (14.88%) | 8 (5.97%) | 26<br>(10.20%) | 0.032 |
| Family History of Hypertension | 48 (39.67%) | 51 (38.06%) | 99<br>(38.82%) | 0.893 |
| History of Pregnancy Induced Hypertension | 16 (17.39%) | 8 (12.50%) | 24<br>(15.38%) | 0.544 |
| 4 or more ANC visit | 43 (35.54%) | 60 (44.78%) | 103<br>(40.39%) | 0.170 |

<sup>a</sup> n (%)
<sup>b</sup> Pearson's Chi-squared test; Fisher's exact test

### 3.3 Maternal outcomes

Maternal mortality did not differ significantly between the groups (3.31% in EO-PE vs. 1.49% in LO-PE, p=0.427). However, specific maternal complications were more frequent in the EO-PE group. PE progressed to eclampsia in 31.40% of EO-PE cases compared to 18.66% in LO-PE cases (p=0.027). Placental abruption was also more common in the EO-PE group (19.83% vs 9.70%, p=0.034). The occurrence of other severe maternal complications such as HELLP syndrome, postpartum hemorrhage (PPH), acute kidney injury (AKI), cerebrovascular accidents or pulmonary edema had no significant differences between the groups. The overall prevalence of adverse maternal outcome (defined as one or more maternal complications or maternal death) was slightly higher in the EO-PE group (57/121, 47.11%) compared to the LO-PE group (60/134, 44.78%). (Table-3)

**Table-3:** Comparison of maternal outcomes between the EO-PE and LO-PE groups.

| Characteristic | Early Onset<br>PE<br>N = 121<sup>a</sup> | Late Onset<br>PE<br>N = 134<sup>a</sup> | Overall<br>N = 255<sup>a</sup> | p-value<sup>b</sup> |
| --- | --- | --- | --- | --- |
| Maternal Mortality | 4 (3.31%) | 2 (1.49%) | 6 (2.35%) | 0.427 |
| Eclampsia | 38 (31.40%) | 25 (18.66%) | 63 (24.71%) | 0.027 |
| Abruptio Placenta | 24 (19.83%) | 13 (9.70%) | 37 (14.51%) | 0.034 |
| HELLP Syndrome | 10 (8.26%) | 10 (7.46%) | 20 (7.84%) | 0.996 |
| Postpartum hemorrhage | 6 (4.96%) | 12 (8.96%) | 18 (7.06%) | 0.213 |
| Premature Rupture of Membrane | 5 (4.13%) | 13 (9.70%) | 18 (7.06%) | 0.136 |
| Acute Kidney Injury | 4 (3.31%) | 2 (1.49%) | 6 (2.35%) | 0.427 |
| Pulmonary Edema | 8 (6.61%) | 4 (2.99%) | 12 (4.71%) | 0.285 |
| Cerebrovascular Accidents | 8 (6.61%) | 8 (5.97%) | 16 (6.27%) | >0.999 |
| Maternal Adverse Outcome | 57 (47.11%) | 60 (44.78%) | 117<br>(45.88%) | 0.805 |

<sup>a</sup> n (%)
<sup>b</sup> Fisher's exact test; Pearson's Chi-squared test

### 3.4 Fetal outcomes

The EO-PE group experienced significantly poorer fetal outcomes. Fetal survival was markedly lower in the EO-PE group (72%; 87 livebirths out of 121 cases) compared to the LO-PE group (97%; 130 livebirths out of 134 cases, p<0.001). Interestingly, women in the LO-PE group had a significantly higher rate of cesarean sections (C/S) compared to those in the EO-PE group (84.33% vs 51.24%, p<0.001). There was no significant difference in sex of the baby between the two groups. (Table-4)

**Table-4:** Comparison of fetal outcomes between the EO-PE and LO-PE groups.

| Characteristic | Early Onset<br>PE<br>N = 121<sup>a</sup> | Late Onset<br>PE<br>N = 134<sup>a</sup> | Overall<br>N = 255<sup>a</sup> | p-value<sup>b</sup> |
| --- | --- | --- | --- | --- |
| **Mode of Delivery** |  |  |  | <0.001 |
| Caesarean Section | 62 (51.24%) | 113 (84.33%) | 175 (68.63%) |  |
| Vaginal Delivery | 59 (48.76%) | 21 (15.67%) | 80 (31.37%) |  |
| **Birth Outcome** |  |  |  | <0.001 |
| Stillbirth | 34 (28.10%) | 4 (2.99%) | 38 (14.90%) |  |
| Livebirth | 87 (71.90%) | 130 (97.01%) | 217 (85.10%) |  |
| **Sex of Baby** |  |  |  | 0.247 |
| Female | 63 (52.07%) | 59 (44.03%) | 122 (47.84%) |  |
| Male | 58 (47.93%) | 75 (55.97%) | 133 (52.16%) |  |

<sup>a</sup> n (%)
<sup>b</sup> Pearson's Chi-squared test; Fisher's exact test

### 3.5 Neonatal outcomes

Among the 217 live births (EO-PE: 87; LO-PE: 130), neonates born to mothers with EO-PE had substantially worse neonatal outcomes compared to those born to mothers with LO-PE. Neonatal death occurred significantly more often in the EO-PE group (11/87, 12.64%) compared to the LO-PE group (2/130, 1.54%). Women in the EO-PE group (75.86%) had significantly higher percentage of premature (<37 weeks) deliveries compared to the LO-PE group (1.54%, p<0.001). Similarly, low birth weight babies (<2500g) were about four times more frequent in the EO-PE group (73.56%) than in the LO-PE group (17.69%, p<0.001). Intrauterine growth retardation (IUGR) was also significantly more common in neonates from the EO-PE group (29.89% vs 14.62%, p=0.011). The difference in the proportion of neonates with a 5-minute Apgar score and the number of cases of congenital anomalies between the EO-PE and LO-PE groups was not significant. The need for neonatal hospital admission was significantly higher for neonates in the EO-PE group compared to the LO-PE group (54.02% vs. 20.77%, p<0.001). Overall, the neonatal adverse outcome was alarmingly higher in the EO-PE group (78/87, 89%) compared to the LO-PE group (33/130, 25.38%). (Table-5)

**Table-5:** Comparison of neonatal outcomes among the livebirths between the EO-PE and LO-PE groups.

| Characteristic | Early Onset<br>PE<br>N = 87<sup>a</sup> | Late Onset<br>PE<br>N = 130<sup>a</sup> | Overall<br>N = 217<sup>a</sup> | p-value<sup>b</sup> |
| --- | --- | --- | --- | --- |
| Neonatal Mortality | 11 (12.64%) | 2 (1.54%) | 13 (5.99%) | 0.002 |
| Prematurity | 66 (75.86%) | 2 (1.54%) | 68 (31.34%) | <0.001 |
| Low Birth Weight | 64 (73.56%) | 23 (17.69%) | 87 (40.09%) | <0.001 |
| Intra Uterine Growth Retardation | 26 (29.89%) | 19 (14.62%) | 45 (20.74%) | 0.011 |
| Congenital Anomaly | 0 (0.00%) | 4 (3.08%) | 4 (1.84%) | 0.151 |
| APGAR score <7 at 5 min | 12 (13.79%) | 10 (7.69%) | 22 (10.14%) | 0.219 |
| Hospital Admission | 47 (54.02%) | 27 (20.77%) | 74 (34.10%) | <0.001 |
| Neonatal Adverse Outcome | 78 (89.66%) | 33 (25.38%) | 111 (51.15%) | <0.001 |

<sup>a</sup> n (%)
<sup>b</sup> Pearson's Chi-squared test; Fisher's exact test

### 3.6 Association between onset of pre-eclampsia and maternal, fetal and neonatal adverse outcomes

In simple logistic regression analyses, EO-PE was not associated with maternal adverse outcomes compared with LO-PE group (OR: 1.10, 95% CI: 0.67–1.80, p=0.7), but was strongly associated with adverse fetal and neonatal outcomes. EO-PE was associated with a markedly increased odds of stillbirth (OR: 12.7, 95% CI: 4.85–43.7, p<0.001). The odds of adverse neonatal outcome was even higher (OR: 25.5, 95% CI: 12.0–59.8, p<0.001).

These associations persisted even after adjusting for maternal age, living area, religion, education, occupation, gravidity, chronic hypertension, hypothyroidism, gestational diabetes mellitus, and ANC attendance. In the multiple logistic regression model, EO-PE remained unassociated with maternal adverse outcomes. However, the odds of adverse fetal outcomes (aOR: 16.3, 95% CI: 5.43–63.2, p<0.001) and adverse neonatal outcomes (aOR: 39.7, 95% CI: 14.7–131, p<0.001) increased significantly in the EO-PE group. Among the individual outcomes, EO-PE was independently associated with higher odds of eclampsia (aOR: 5.08, 95% CI: 2.24–12.36; p < 0.001), abruptio placenta (aOR: 2.56, 95% CI: 1.08–6.31; p = 0.032), and low birth weight (aOR: 32.60, 95% CI: 11.68–109.38; p < 0.001). No significant independent associations were observed for HELLP syndrome (aOR: 1.41, 95% CI: 0.47–4.41; p = 0.544) or intrauterine growth retardation (aOR: 1.84, 95% CI: 0.75–4.59; p = 0.183). (Figure-2)

**Figure-2:**
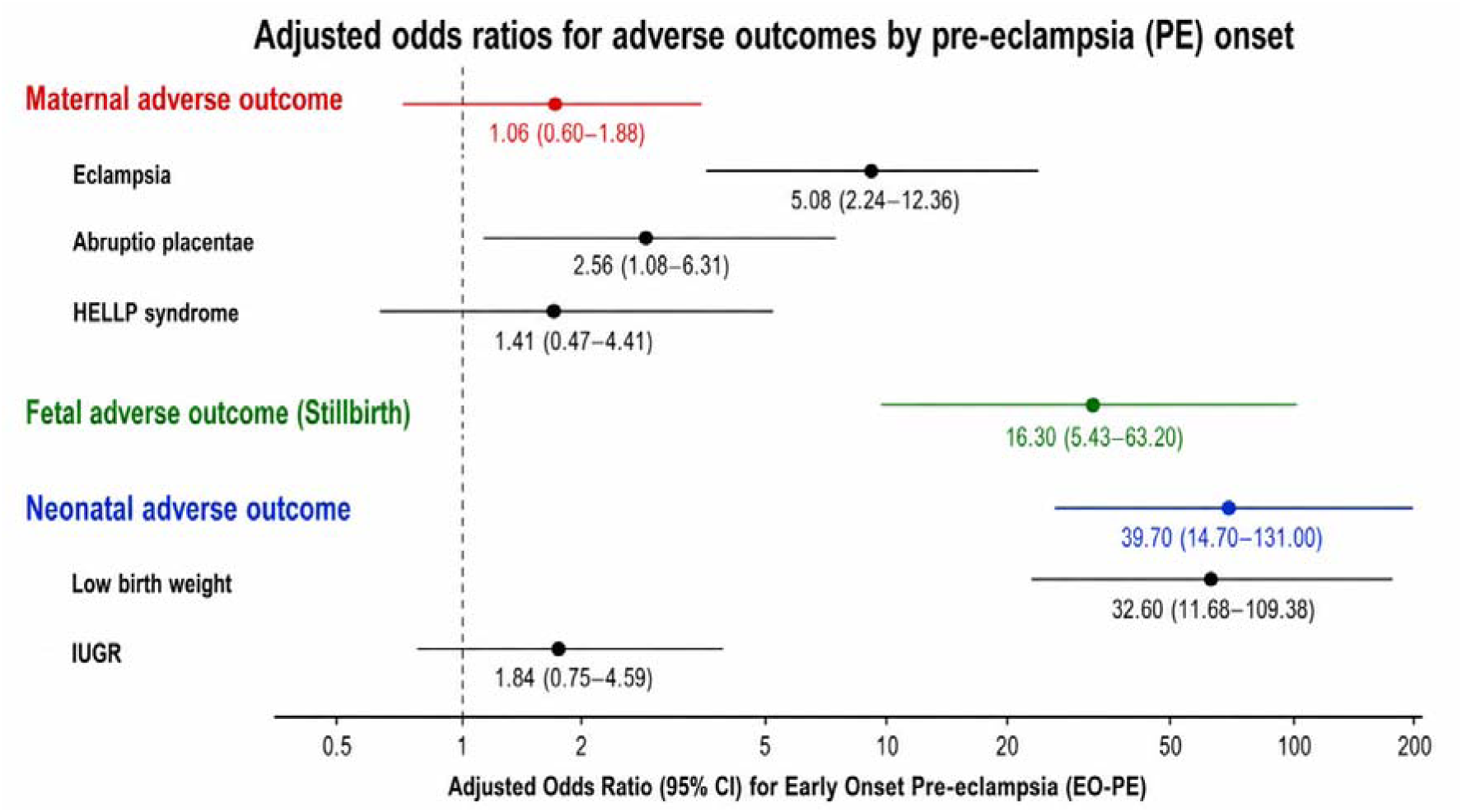
Adjusted association between pre-eclampsia onset and adverse maternal, fetal and neonatal outcomes

## 4. Discussion

This retrospective cohort study showed that early-onset PE was associated with more severe maternal complications such as eclampsia and placental abruption, and substantially worse fetal and neonatal adverse outcomes including higher rates of stillbirth, prematurity, low birth weight and neonatal mortality. Our findings align with previous research suggesting that early-onset PE (EO-PE) represents a more severe form of the disorder (12–15).

The observed prevalence of PE (11.28%) among admitted pregnant women in our study is consistent with reports from some other studies in the region. Recent facility-based studies in Nepal and Bangladesh reported the prevalence of PE as 12% and 14% respectively (16, 17). This high prevalence in tertiary centers is likely indicative of referral bias, where complicated cases are disproportionately represented. Such tertiary centers should be adequately resourced with specialized personnel and infrastructure to handle the high volume of these patients.

Studies have consistently shown that women with EO-PE have a higher incidence of severe maternal complications (18–20). In our cohort, EO-PE was associated with significantly higher rates of eclampsia and placental abruption, consistent with findings from comparable settings in India (21). The underlying cause of these poor outcomes lies in the defective placentation that occurs early in pregnancy, leading to inadequate spiral artery remodeling, uteroplacental ischemia, and a surge of anti-angiogenic factors into the maternal bloodstream (13, 22). These factors contribute to widespread endothelial dysfunction. Given its severity, EO-PE should be treated as a high-risk condition that requires close monitoring and care in specialized settings. Improving early screening and educating women about warning signs during pregnancy could help reduce serious maternal complications.

The significantly higher caesarean section rate in the LO-PE group (84.33% vs. 51.24%) deserves careful interpretation. In EO-PE, expectant management at <34 weeks is clinically rational to allow fetal maturation, often resulting in more vaginal deliveries. By contrast, in LO-PE at ≥34 weeks, planned caesarean delivery is more commonly initiated electively. This pattern likely reflects local institutional protocols rather than a genuine difference in severity-driven operative delivery.

This study reported that EO-PE was associated with poorer fetal survival and increased rates of prematurity, low birth weight, IUGR and neonatal death. Our multivariable analysis demonstrated substantially higher odds of adverse fetal and neonatal outcomes among EO-PE cases, independent of maternal sociodemographic characteristics. These adverse perinatal outcomes reflect the severe uteroplacental insufficiency characteristic of EO-PE, where the placenta fails to develop properly early in pregnancy (13). As a result, it cannot provide adequate oxygen and nutrients to the fetus (23). This dysfunctional placenta poses serious risks to both the baby and the mother. To protect the mother’s health, doctors often need to deliver the baby prematurely through iatrogenic preterm birth. While this can be life-saving for the mother, it results in neonatal prematurity and its associated complications such as breathing difficulties, feeding problems, and long-term disability (24, 25). This pathophysiology justifies the higher rate of hospitalization and neonatal mortality among children born to mothers with EO-PE, which was also observed in our study. Management of these cases must occur exclusively in integrated tertiary centers with a Neonatal Intensive Care Unit (NICU). Additionally, specialized training should be facilitated for healthcare providers for a better maternal and neonatal outcome.

### 4.1 Clinical and public health implications

The findings of this study carry direct and actionable implications for clinical practice in Bangladesh and comparable low-resource settings. PE onset timing should be formally incorporated into clinical risk stratification at the point of diagnosis. Women with EO-PE should be managed exclusively in NICU-equipped tertiary centres with standardised protocols for magnesium sulphate prophylaxis, antihypertensive therapy, antenatal corticosteroid administration, and timely delivery planning. First-trimester PE risk screening using the FIGO combined algorithm, followed by low-dose aspirin prophylaxis for high-risk women identified before 16 weeks, offers a feasible preventive strategy that warrants integration into national ANC guideline (26, 27). Public health efforts should prioritize strengthening referral systems, expanding NICU services in regional hospitals, and enhancing the skills of frontline maternity care providers.

### 4.2 Strengths and limitations

To the best of our knowledge, this is the first study from Bangladesh to compare pregnancy outcomes based on the timing of pre-eclampsia onset. An additional strength lies in the comprehensive evaluation of maternal, fetal, and neonatal outcomes, enabling a multidimensional understanding of the clinical burden associated with PE. Several limitations must be acknowledged. First, the retrospective cohort design relies on the completeness and accuracy of hospital records which may have resulted in outcome misclassification. Second, the single tertiary referral centre design introduces referral bias as it represents complicated PE cases referred from lower-level facilities. Third, the multivariable models did not include key treatment variables including magnesium sulphate use, antihypertensive regimen, and antenatal corticosteroid administration, Additionally, important confounders such as referral status and gestational age at delivery were also unavailable. Fourth, the collection of sociodemographic and clinical history variables via telephone interview introduces potential recall bias. Fifth, the high percentage of exclusion rate may have led to selection bias, although comparison of available characteristics between included and excluded patients revealed no statistically significant differences. Sixth, the study period of six months precludes assessment of long-term maternal or neonatal complications of pre-eclampsia.

### 4.3 Future research directions

Future research should address the limitations of this study and extend its findings in several directions. Most urgently, multicentre prospective cohort studies across geographically diverse facilities in Bangladesh are needed to generate nationally representative, onset-stratified PE outcome data. Such studies should incorporate formal disease severity classification, complete treatment variable documentation, and placental pathology reporting, enabling a more granular understanding of which clinical factors mediate the excess risk observed in EO-PE. Longitudinal follow-up of neonates born to mothers with EO-PE is essential to quantify the full burden of EO-PE exposure on child health in a low-resource context. Developing and externally validating prediction models integrating maternal biomarkers, uterine artery Doppler indices, and clinical risk factors should be prioritized to facilitate earlier identification of women at high risk of EO-PE. Finally, implementation research evaluating the feasibility, acceptability, and cost-effectiveness of integrating first-trimester PE risk screening and aspirin prophylaxis into Bangladesh’s national ANC protocol is urgently needed to translate the evidence base into policy and practice.

## 5. Conclusion

This retrospective cohort study from Bangladesh’s first onset-stratified PE analysis demonstrates that EO-PE carries a substantially higher burden of adverse perinatal outcomes including stillbirth, prematurity, low birth weight, and neonatal mortality compared with LO-PE. These findings identify women with EO-PE as a high-risk clinical subgroup who require timely referral, intensive monitoring, and management in tertiary care facilities with adequate maternal and neonatal critical care capacity. From a public health perspective, integrating first-trimester preeclampsia risk assessment into national antenatal care guidelines, alongside strengthening referral systems and regional neonatal intensive care services, may help reduce the burden of EO-PE. Future multicentre prospective studies incorporating long-term maternal and neonatal follow-up, and externally validated prediction models are needed to guide targeted interventions in Bangladesh and other low-resource settings.

## Data Availability

All data produced are available online in Mendeley Data at the following link:

http://doi.org/10.17632/t837f3g2kd.1

## ACKNOWLEDGEMENTS

The authors gratefully acknowledge the Department of Obstetrics & Gynaecology at Faridpur Medical College Hospital (FMCH) for their invaluable support in providing patient records for this study.

## REFERENCES

1. Poon LC, Shennan A, Hyett JA, Kapur A, Hadar E, Divakar H, et al. The International Federation of Gynecology and Obstetrics (FIGO) initiative on preeclampsia (PE): a pragmatic guide for first trimester screening and prevention. International journal of gynaecology and obstetrics: the official organ of the International Federation of Gynaecology and Obstetrics. 2019;145(Suppl 1):1.

2. Duckitt K, Harrington D. Risk factors for pre-eclampsia at antenatal booking: systematic review of controlled studies. BMJ. 2005;330(7491):565.

3. Obstetricians ACo, Gynecologists. Hypertension in pregnancy. Report of the American College of Obstetricians and Gynecologists’ task force on hypertension in pregnancy. Obstet Gynecol. 2013;122:1122.

4. Kharaghani R, Cheraghi Z, Esfahani BO, Mohammadian Z, Nooreldinc RS. Prevalence of preeclampsia and eclampsia in Iran. Arch Iran Med. 2016;19(1):0-.

5. Basso O, Rasmussen S, Weinberg CR, Wilcox AJ, Irgens LM, Skjaerven R. Trends in fetal and infant survival following preeclampsia. JAMA. 2006;296(11):1357–62.

6. Osungbade KO, Ige OK. Public health perspectives of preeclampsia in developing countries: implication for health system strengthening. Journal of pregnancy. 2011;2011(1):481095.

7. Ali A, Islam J, Paul R, Parvin S, Mohammed Mohiuddin Chowdhury AT, Islam R, et al. Geographic inequalities and determinants of anaemia among preeclamptic women: a cross-sectional sample-based study in Bangladesh. BMC Public Health. 2024;24(1):1650.

8. Khan S, Siddique AB, Jabeen S, Hossain AT, Haider MM, Zohora FT, et al. Preeclampsia and eclampsia-specific maternal mortality in Bangladesh: Levels, trends, timing, and care-seeking practices. Journal of Global Health. 2023;13:07003.

9. Rahman S, Begum N, Mahmud A. Evaluation of Morbidity and Mortality of Eclampsia at Brahmanbaria Medical College and Hospital. Journal of Rangpur Medical College. 2024;9(1):39–43.

10. Von Dadelszen P, Magee LA, Roberts JM. Subclassification of preeclampsia. Hypertens Pregnancy. 2003;22(2):143–8.

11. Roberts CL, Ford JB, Algert CS, Antonsen S, Chalmers J, Cnattingius S, et al. Population-based trends in pregnancy hypertension and pre-eclampsia: an international comparative study. BMJ Open. 2011;1(1):e000101.

12. Raymond D, Peterson E. A critical review of early-onset and late-onset preeclampsia. Obstet Gynecol Surv. 2011;66(8):497–506.

13. Redman C, Sargent I. Placental stress and pre-eclampsia: a revised view. Placenta. 2009;30:38–42.

14. Valensise H, Vasapollo B, Gagliardi G, Novelli GP. Early and late preeclampsia: two different maternal hemodynamic states in the latent phase of the disease. Hypertension. 2008;52(5):873–80.

15. LaMarca BD, Gilbert J, Granger JP. Recent progress toward the understanding of the pathophysiology of hypertension during preeclampsia. Hypertension. 2008;51(4):982–8.

16. Shrestha J, Subedi A, Gauchan E, Shrestha A, Pandey C. Pregnancy outcome in early versus late onset preeclampsia. Nepal Journal of Obstetrics and Gynaecology. 2021;16(2):53–9.

17. Mou AD, Barman Z, Hasan M, Miah R, Hafsa JM, Das Trisha A, Ali N. Prevalence of preeclampsia and the associated risk factors among pregnant women in Bangladesh. Sci Rep. 2021;11(1):21339.

18. Wojtowicz A, Zembala-Szczerba M, Babczyk D, Kołodziejczyk-Pietruszka M, Lewaczyńska O, Huras H. Early-and late-onset preeclampsia: a comprehensive cohort study of laboratory and clinical findings according to the new ISHHP criteria. Int J Hypertens. 2019;2019(1):4108271.

19. Wadhwani P, Saha PK, Kalra JK, Gainder S, Sundaram V. A study to compare maternal and perinatal outcome in early vs. late onset preeclampsia. Obstetrics & Gynecology Science. 2020;63(3):270.

20. Lisonkova S, Joseph K. Incidence of preeclampsia: risk factors and outcomes associated with early-versus late-onset disease. Am J Obstet Gynecol. 2013;209(6):544. e1-. e12.

21. Geeta Sri LK, Lakshmi CS. Early and Late Onset Pre-eclampsia – Maternal and Perinatal Outcome in Tertiary Care Hospital. International Journal of Current Microbiology and Applied Sciences. 2021;10(8):156–63.

22. Rana S, Lemoine E, Granger JP, Karumanchi SA. Preeclampsia: pathophysiology, challenges, and perspectives. Circ Res. 2019;124(7):1094–112.

23. Burton GJ, Redman CW, Roberts JM, Moffett A. Pre-eclampsia: pathophysiology and clinical implications. BMJ. 2019;366.

24. Madazli R, Yuksel MA, Imamoglu M, Tuten A, Oncul M, Aydin B, Demirayak G. Comparison of clinical and perinatal outcomes in early-and late-onset preeclampsia. Arch Gynecol Obstet. 2014;290:53–7.

25. Schneider H. Placental dysfunction as a key element in the pathogenesis of preeclampsia. Journal of Mother and Child. 2017;21(4):309–16.

26. Rolnik DL, Wright D, Poon LC, O’Gorman N, Syngelaki A, de Paco Matallana C, et al. Aspirin versus placebo in pregnancies at high risk for preterm preeclampsia. N Engl J Med. 2017;377(7):613–22.

27. Roberge S, Bujold E, Nicolaides KH. Aspirin for the prevention of preterm and term preeclampsia: systematic review and metaanalysis. Am J Obstet Gynecol. 2018;218(3):287–93. e1.

